# Beyond Nodal Status: Interactions Between Molecular Subtype, Tumor Burden, and Survival in 12,225 Patients with Breast Cancer

**DOI:** 10.64898/2026.06.22.26356207

**Authors:** Majid Akrami, Nastaran Tavakolian, Hooman Arianpour, Amirhesam Moosazadeh, Amir Hossein Rajabi, Zahra Keumarsi, Masoumeh Ghoddusi Johari, Vahid Zangouri, Abdolrasoul Talei

## Abstract

**Background:** Lymph node status and molecular subtype are among the most established prognostic factors in breast cancer. However, the extent to which their prognostic effects vary across different tumor size categories and clinical subgroups remains incompletely understood. We investigated the interplay between nodal status, molecular subtype, and tumor size in a large real world breast cancer cohort and developed a prognostic nomogram for individualized survival prediction.

**Methods:** A total of 12,225 women with invasive breast cancer from the Shiraz Breast Cancer Registry were analyzed. Patients were stratified according to tumor size, lymph node status, and molecular subtype. Overall survival (OS) and disease free survival (DFS) were evaluated using Kaplan Meier analyses and subgroup comparisons. Logistic regression was performed to identify predictors of lymph node involvement, while Cox regression was used to determine independent prognostic factors. A nomogram was subsequently developed and internally validated for prediction of 3-year and 5-year OS.

**Results:** Of 12,225 patients, 41.7% had lymph node positive disease. Across nearly all tumor size categories and molecular subtypes, nodal involvement was associated with significantly worse OS and DFS. Notably, the survival disadvantage associated with nodal positivity was more pronounced among patients with larger tumors and among those with HER2 positive and triple negative breast cancer (TNBC). Although TNBC demonstrated the lowest rate of lymph node involvement among molecular subtypes (adjusted OR 0.54, 95% CI 0.46-0.63), it appeared to show one of the largest survival gaps between node positive and node negative disease. In the overall cohort, survival outcomes generally ranked from best to worst as Luminal A, Luminal B, HER2 positive, and TNBC. However, survival differences among molecular subtypes were not consistently observed across all tumor size and nodal status subgroups. When significant differences were present, Luminal A and Luminal B tumors consistently showed superior outcomes compared with HER2 positive and TNBC tumors. Multivariable analysis identified lymph node status, tumor size, molecular subtype, lymphovascular invasion, tumor necrosis, type of surgery, radiotherapy, hormone therapy, and adjuvant chemotherapy as independent prognostic factors. A nomogram integrating clinicopathological and treatment variables demonstrated good predictive performance, with time dependent AUCs of 0.749 and 0.751 for 3 year and 5 year OS, respectively, and showed good calibration.

**Conclusions:** The prognostic impact of lymph node status is not uniform across breast cancer subgroups and appears particularly pronounced in larger tumors and biologically aggressive subtypes. Despite a lower likelihood of nodal involvement, TNBC showed substantial outcome deterioration when nodal metastasis was present. These findings highlight the importance of jointly considering nodal status, molecular subtype, and tumor burden in prognostic assessment.

## Introduction

Breast cancer (BC) is the most frequently diagnosed cancer among women in 157 countries and the leading cause of cancer-related deaths in 112 countries. In 2022, there were approximately 2.3 million new cases of female breast cancer reported worldwide, leading to 666,000 deaths attributed to the disease. It accounts for about one in four of all diagnosed cancers and one in six cancer-related deaths in women (1).

The introduction of gene expression DNA microarrays provided a new method for classifying breast cancer in the early 2000s (2). By analyzing the expression levels of thousands of genes in tumors, distinct groups with similar patterns termed intrinsic or molecular subtypes were identified, namely luminal A, luminal B, HER2-positive, and triple negative. These molecular subtypes impact prognosis and treatment, affecting survival outcomes and the choice of targeted therapies. (3, 4).

Over the past few decades, the prognosis and survival for breast cancer patients have significantly improved. Between 1989 and 2022, breast cancer mortality rates decreased by 44%, which has saved approximately 518,000 lives in the United States. These advancements can be attributed to earlier detection by screening and progress in treatment protocols (5).

Modern breast cancer therapies have significantly evolved due to a deeper understanding of molecular diversity, resulting in more targeted treatment options. Advancements in treatment have improved cure rates for early-stage breast cancer to about 70–80%. However, for those with advanced breast cancer, which remains classified as incurable, the focus has shifted to prolonging survival, managing symptoms, and improving the quality of life for patients (6).

Numerous studies conducted over the years have highlighted several prognostic factors in breast cancer, such as patient age, lymph node status, tumor dimensions, histopathological grade, as well as molecular markers like estrogen receptor (ER) status, progesterone receptor (PR) status, human epidermal growth factor receptor 2 ) HER2( expression, and Ki-67 levels (7–9).

Lymph node status is one of the most significant prognostic factors in breast cancer. It plays a crucial role in tumor staging, as defined by the American Joint Committee on Cancer (AJCC) system. This staging system considers three key factors: tumor size (T), lymph node status (N), and distant metastasis (M). Together, these factors help determine the stage of the disease and guide treatment strategies (10–12).

The development of the pre-metastatic lymph node niche is driven by multiple cellular and molecular mechanisms. These include lymph-angiogenesis, extracellular matrix remodeling, and the recruitment of immunosuppressive cell populations such as macrophages and regulatory T cells (13). In parallel with nodal invasion, the frequency of Tregs increases. These cells also exhibit higher expression levels of co-inhibitory and co-stimulatory receptors than effector immune cells, resulting in significant alterations within the immune microenvironment (14).

In this study, we analyzed a large cohort of 12,225 women with breast cancer from southern Iran. Patients were stratified into different groups based on tumor size, lymph node involvement, and molecular subtype. Survival analyses were then performed to compare prognostic outcomes of different molecular subtypes and lymph node status across these groups. Independent prognostic factors of breast cancer were identified, and finally, a nomogram incorporating patient, tumor, and treatment characteristics was developed to predict patient survival.

## Materials and methods

### Study Design and Data Source

This study utilized data from the Shiraz Breast Cancer Registry (SBCR), one of the largest institutional breast cancer registries in southern Iran. The registry includes patients who underwent surgical treatment for primary breast cancer at affiliated centers. Data collection began in 2000 and includes prospective follow-up for recurrence and survival. For the present study, all eligible female patients diagnosed with breast cancer were included and followed until September 2024.

### Inclusion and Exclusion Criteria

Patients were included if they had histologically confirmed invasive breast carcinoma and underwent definitive surgery (either breast-conserving surgery or mastectomy). Exclusion criteria included the presence of distant metastasis at diagnosis (stage IV), male patients with breast cancer, missing lymph node status, or incomplete records for key clinicopathological variables.

### Variables and Definitions

The variables used in this study included both patient-related and tumor-related characteristics. Patient-related variables were the year of diagnosis, age at diagnosis, type of surgery, and whether the patient received radiotherapy, chemotherapy (either neoadjuvant or adjuvant), or endocrine therapy.

Tumor characteristics were described according to pathology reports. Tumor size was categorized as less than 2 cm, between 2 and 5 cm, or greater than 5 cm, based on the maximum pathological dimension and corresponding to AJCC T1–T3 categories. Lymph node status was considered positive if at least one axillary lymph node contained metastatic involvement. Tumor grade was assigned using standard histological grading, and the presence or absence of tumor necrosis was determined based on pathology findings. Lymphovascular invasion (LVI) was considered present if there was any indication of lymphatic or blood vessel involvement.

Tumors were categorized as Luminal A, Luminal B, HER2-positive, and triple-negative breast cancer (TNBC).

Recurrence was categorized as local/regional or metastatic. Overall survival (OS) was defined as the time from diagnosis to death from any cause, and disease-free survival (DFS) as the time from primary treatment to recurrence or death. Patients who had not experienced the event of interest, either death or recurrence, by the end of the study period or who were lost to follow-up were considered censored.

### Descriptive Statistics and Group Comparisons

Normally distributed continuous variables were presented as mean ± standard deviation (SD), while categorical variables were summarized using counts and percentages. Group differences between LN+ and LN− were assessed using the chi-squared test or Fisher’s exact test, as appropriate.

### Survival Analysis

Kaplan–Meier survival curves were generated for 5-year OS and 5-year DFS by lymph node status, tumor size, and molecular subtype. Survival distributions were compared using the log-rank test. Stratified subgroup analyses were conducted to assess survival differences within each molecular subtype and tumor size category.

To identify predictors of lymph node involvement, univariate and multivariate logistic regression models were constructed. Based on significance, Candidate predictors included tumor size, lymphovascular invasion, tumor necrosis, and molecular subtype. Additional variables (e.g., tumor grade, multifocality, age) were considered but excluded due to multicollinearity or high missingness. Adjusted odds ratios (ORs) with 95% confidence intervals (CIs) were reported.

### Univariate and Multivariate Analysis

Cox univariate and multivariate regression models were used to identify factors affecting patient survival. Proportional hazards assumptions were assessed visually and statistically using Schoenfeld residuals where applicable. Multicollinearity was assessed. Adjusted hazard ratios (HRs) with 95% CIs were reported.

### Nomogram Development and Validation

To enhance the clinical utility of the findings, a prognostic nomogram was developed to estimate 3-year and 5-year overall survival probabilities. The model incorporated nine clinically and statistically significant predictors: type of surgery, molecular subtype, tumor necrosis, lymph node status, tumor size, lymphovascular invasion, adjuvant chemotherapy, radiotherapy, and hormone therapy. The nomogram was constructed using the *rms* and *ggplot2* packages in R. Model discrimination was assessed using the concordance index (C-index), indicating predictive accuracy. Time-dependent receiver operating characteristic (ROC) curves were generated to further evaluate model performance. Model calibration was assessed using calibration plots at 3 and 5 years, which demonstrated agreement between predicted and observed survival probabilities across risk strata.

### Statistical Analysis

All statistical analyses were conducted using SPSS version 27.0 (IBM Corp., Armonk, NY) and R version 4.5. A two-tailed p-value of <0.05 was considered statistically significant.

## Results

### Patient Characteristics and Clinicopathological Features

Of the 12,225 women included in the Shiraz Breast Cancer Registry, 5,097 (41.7%) were lymph node-positive (LN+) and 7,128 (58.3%) were lymph node-negative (LN−). Left breast involvement was slightly more common in both groups, although the difference in tumor laterality was not statistically significant (p = 0.734).

Breast-conserving surgery was performed more frequently in the LN− group (60.3%) compared to the LN+ group, where mastectomy was the predominant procedure (52.1%; p < 0.001).

All major tumor characteristics differed significantly between the two groups (all p < 0.001). Most patients in both groups had T2 tumors (2–5 cm), with a higher proportion in the LN+ group (65.2% vs. 57.6%). Grade II tumors were the most common histological grade, observed in 63.0% of LN+ and 56.7% of LN− patients. The luminal A subtype was predominant in both cohorts (60.0% in LN+ vs. 56.9% in LN−). Lymphovascular invasion (LVI) was present in 64.9% of LN+ patients, whereas 81.4% of LN− patients were LVI-negative. Multifocality was uncommon in both groups (>90% solitary tumors). Tumor necrosis was more frequent in the LN+ group (46.7% vs. 30.9%). Clear surgical margins were achieved in the majority of cases, though slightly less often in LN+ patients (95.9% vs. 97.7%; p < 0.001).

Significant differences were also observed in adjuvant treatments received (all p < 0.001). Hormone therapy and radiotherapy were administered more frequently to LN+ patients (70.7% and 78.9%, respectively) than to LN− patients (57.3% and 52.4%, respectively). Intraoperative radiotherapy (IORT) was rarely used in either group. Adjuvant chemotherapy was given to over 69.1% of patients overall and was significantly more common in the LN+ group (81.1% vs. 60.5%).

Tumor recurrence occurred in 25.8% of LN+ patients compared with 14.1% of LN− patients (p < 0.001). Distant metastasis was the most common pattern of recurrence in both groups. At the end of follow-up, 85.8% of the entire cohort remained alive, with significantly lower survival in the LN+ group (80.9% vs. 89.7%; p < 0.001). Detailed clinicopathological characteristics are summarized in Table 1.

**Table 1.** Baseline clinicopathological characteristics of patients.

| Characteristics | Lymph node status |  | Total | P Value |
| --- | --- | --- | --- | --- |
|  | Positive | Negative |  |  |
| Breast laterality |  |  |  |  |
| Right | 2444(48.0) | 2995(48.3) | 5439(48.2) | 0.734 |
| Left | 2650(52.0) | 3206(51.7) | 5856(51.8) |  |
| Type of surgery |  |  |  |  |
| BCS | 2434(47.9) | 3680(60.3) | 6114(54.7) | <0.001 |
| Mastectomy | 2646(52.1) | 2426(39.7) | 5072(45.3) |  |
| Tumor size |  |  |  |  |
| T1(≤2 cm) | 1179(25.0) | 1772(36.5) | 2951(30.9) | <0.001 |
| T2(2-5 cm) | 3068(65.2) | 2791(57.6) | 5859(61.3) |  |
| T3(>5cm) | 461(9.8) | 285(5.9) | 746(7.8) |  |
| Tumor grade |  |  |  |  |
| I | 742(16.5) | 953(22.2) | 1695(19.3) | <0.001 |
| II | 2824(63.0) | 2429(56.7) | 5253(59.9) |  |
| III | 920(20.5) | 907(21.1) | 1827(20.8) |  |
| Molecular subtypes |  |  |  |  |
| Luminal A | 2509(60.0) | 2479(56.9) | 4988(58.4) | <0.001 |
| Luminal B | 808(19.3) | 723(16.6) | 1531(17.9) |  |
| Her2-enriched | 466(11.1) | 451(10.3) | 917(10.7) |  |
| Triple negative | 399(9.5) | 706(16.2) | 1105(12.9) |  |
| Lymphovascular invasion (LVI) |  |  |  |  |
| Yes | 3311(64.9) | 1327(18.6) | 4638(37.9) | <0.001 |
| No | 1786(35.1) | 5801(81.4) | 7587(62.1) |  |
| Multifocality |  |  |  |  |
| Yes | 409(8.0) | 249(3.5) | 658(5.4) | <0.001 |
| No | 4688(92.0) | 6879(96.5) | 11567(94.6) |  |
| In situ component |  |  |  |  |
| Yes | 2171(42.6) | 4435(62.2) | 6606(54.0) | <0.001 |
| No | 2926(57.4) | 2693(37.8) | 5619(46.0) |  |
| Tumor necrosis |  |  |  |  |
| Yes | 2381(46.7) | 2200(30.9) | 4581(37.5) | <0.001 |
| No | 2716(53.3) | 4928(69.1) | 7644(62.5) |  |
| Surgical margin status |  |  |  |  |
| Free | 4888(95.9) | 6962(97.7) | 11850(96.9) | <0.001 |
| Close | 13(0.3) | 14(0.2) | 27(0.2) |  |
| Positive | 196(3.8) | 152(2.1) | 348(2.8) |  |
|  | Positive | Negative |  |  |
| Hormone therapy |  |  |  |  |
| Yes | 3605(70.7) | 4083(57.3) | 7688(62.9) | <0.001 |
| No | 1492(29.3) | 3045(42.7) | 4537(37.1) |  |
| IORT |  |  |  |  |
| Yes | 5042(98.9) | 6768(94.9) | 11810(96.6) | <0.001 |
| No | 55(1.1) | 360(5.1) | 415(3.4) |  |
| Radiotherapy |  |  |  |  |
| Yes | 4019(78.9) | 3732(52.4) | 7751(63.4) | <0.001 |
| No | 1078(21.1) | 3396(47.6) | 4474(36.6) |  |
| Neoadjuvant chemotherapy |  |  |  |  |
| Yes | 977(19.2) | 893(12.5) | 1870(15.3) | <0.001 |
| No | 4120(80.8) | 6235(87.5) | 10355(84.7) |  |
| Adjuvant chemotherapy |  |  |  |  |
| Yes | 4134(81.1) | 4311(60.5) | 8445(69.1) | <0.001 |
| No | 963(18.9) | 2817(39.5) | 3780(30.9) |  |
| Tumor recurrence |  |  |  |  |
| Yes | 1288(25.8) | 879(14.1) | 2167(19.3) | <0.001 |
| No | 3699(74.2) | 5370(85.9) | 9069(80.7) |  |
| Recurrence pattern |  |  |  |  |
| Local | 154(12.1) | 179(20.7) | 333(15.6) | <0.001 |
| Metastatic | 1117(87.9) | 685(79.3) | 1802(84.4) |  |
| Survival status |  |  |  |  |
| Alive | 4033(80.9) | 5604(89.7) | 9637(85.8) | <0.001 |
| Dead | 954(19.1) | 645(10.3) | 1599(14.2) |  |

## Logistic Regression Analysis for Predictors of Lymph Node Involvement

Table 2 summarizes the univariable and multivariable logistic regression analyses for predictors of lymph node involvement.

**Table 2.** Univariable and multivariable logistic regression analyses examining factors associated with lymph node involvement.

| Variables | Univariable OR (95% CI) | *P* Value | Multivariable OR (95% CI) | *P* Value |
| --- | --- | --- | --- | --- |
| Tumor size |  |  |  |  |
| T1(≤2 cm) | Reference | -- | Reference | -- |
| T2(2-5 cm) | 1.65(1.51 to 1.80) | < 0.001 | 1.43(1.28 to 1.59) | < 0.001 |
| T3(>5cm) | 2.431(2.06 to 2.86) | < 0.001 | 2.35(1.90 to 2.90) | < 0.001 |
| Lymphovascular invasion (LVI) |  |  |  |  |
| Absent | Reference | -- | Reference | -- |
| Present | 8.10(7.46 to 8.81) | < 0.001 | 5.55(5.07 to 6.13) | < 0.001 |
| Tumor necrosis |  |  |  |  |
| Absent | Reference | -- | Reference | -- |
| Present | 1.96(1.82 to 2.11) | < 0.001 | 1.03(0.93 to 1.14) | 0.55 |
| Molecular subtypes |  |  |  |  |
| Luminal A | Reference | -- | Reference | -- |
| Luminal B | 1.10(0.98 to 1.23) | 0.09 | 1.04(0.91 to 1.20) | 0.48 |
| Her2-enriched | 1.02(0.88 to 1.17) | 0.77 | 0.88(0.75 to 1.05) | 0.16 |
| Triple negative | 0.55(0.48 to 0.63) | <0.001 | 0.54(0.46 to 0.63) | <0.001 |

Tumor size was significantly associated with lymph node positivity. Compared with T1 tumors (≤2 cm), T2 tumors (2–5 cm) were associated with higher odds of lymph node involvement in both univariable (OR 1.65, 95% CI 1.51–1.80; p < 0.001) and multivariable analyses (OR 1.43, 95% CI 1.28–1.59; p < 0.001). Similarly, T3 tumors (>5 cm) showed significantly increased odds relative to T1 tumors in univariable (OR 2.43, 95% CI 2.06–2.86; p < 0.001) and multivariable models (OR 2.35, 95% CI 1.90–2.90; p < 0.001).

Lymphovascular invasion (LVI) was the strongest predictor of lymph node involvement. In univariable analysis, the presence of LVI increased the odds more than eightfold (OR 8.10, 95% CI 7.46–8.81; p < 0.001). This association remained robust after multivariable adjustment (OR 5.55, 95% CI 5.07–6.13; p < 0.001).

Tumor necrosis was significantly associated with lymph node involvement in univariable analysis (OR 1.96, 95% CI 1.82–2.11; p < 0.001); however, this association was no longer significant after adjustment for other covariates (OR 1.03, 95% CI 0.93–1.14; p = 0.55).

About molecular subtypes (reference: luminal A), triple-negative breast cancer was associated with significantly lower odds of lymph node involvement in both univariable (OR 0.55, 95% CI 0.48–0.63; p < 0.001) and multivariable analyses (OR 0.54, 95% CI 0.46–0.63; p < 0.001). Luminal B tumors showed a marginally increased odds in univariable analysis (OR 1.10, 95% CI 0.98–1.23; p = 0.09) that did not persist after multivariable adjustment (OR 1.04, 95% CI 0.91–1.20; p = 0.48). HER2-enriched tumors were not significantly associated with lymph node involvement in either univariable (OR 1.02, 95% CI 0.88–1.17; p = 0.77) or multivariable analysis (OR 0.88, 95% CI 0.75–1.05; p = 0.16).

### Survival Analyses in Subgroups by Lymph Node Status

As shown in Table 3, five-year overall survival differed significantly between LN+ and LN− patients across all tumor size categories (p < 0.001 for T1 and T2; p = 0.003 for T3). In LN+ patients, 5-year OS rates were 90% for T1 tumors (≤2 cm), 89% for T2 (2–5 cm), and 76% for T3 tumors (>5 cm) (Fig. 1a). Corresponding rates in LN− patients were 95% for both T1 and T2 tumors and 87% for T3 tumors (Fig. 1b).

**Figure 1.**
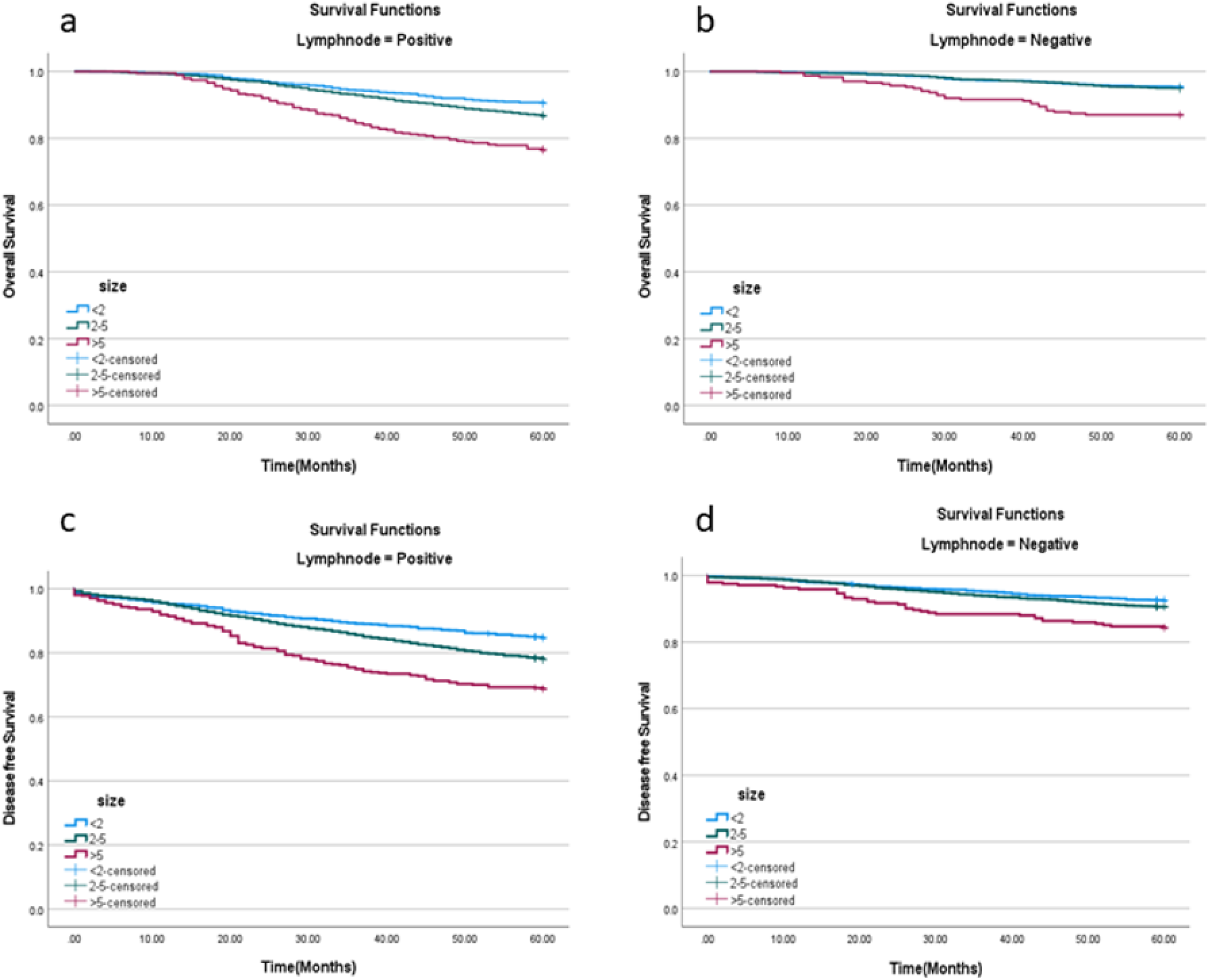
Kaplan–Meier curves for overall survival in LN+ (a) and LN− (b) patients, and disease-free survival in LN+ (c) and LN− (d) patients, stratified by tumor size (T stage).

**Table 3.** Five-year overall survival (OS) and disease-free survival (DFS) stratified by molecular subtype, tumor size, and recurrence type in patients with positive and negative lymph node status.

|  | **Lymph node status** |  |  |
| --- | --- | --- | --- |
|  | **Positive** | **Negative** | **P Value** |
| **Overall survival** |  |  |  |
| Tumor size |  |  |  |
| T1(≤2 cm) | 90% | 95% | <0.001 |
| T2(2-5 cm) | 89% | 95% | <0.001 |
| T3(>5cm) | 76% | 87% | 0.003 |
| Molecular subtypes |  |  |  |
| Luminal A | 89% | 95% | <0.001 |
| Luminal B | 87% | 94% | <0.001 |
| Her2-enrichd | 80% | 92% | <0.001 |
| Triple negative | 76% | 90% | <0.001 |
| Recurrence pattern |  |  |  |
| Local | 83% | 88% | 0.224 |
| Metastatic | 58% | 62% | 0.013 |
| **Disease free survival** |  |  |  |
| Tumor size |  |  |  |
| T1(≤2 cm) | 84% | 92% | <0.001 |
| T2(2-5 cm) | 77% | 91% | <0.001 |
| T3(>5cm) | 69% | 84% | <0.001 |
| Molecular subtypes |  |  |  |
| Luminal A | 80% | 91% | <0.001 |
| Luminal B | 80% | 90% | <0.001 |
| Her2-enrichd | 70% | 86% | <0.001 |
| Triple negative | 70% | 87% | <0.001 |

A comparable pattern was observed for five-year disease-free survival, with significant differences between LN+ and LN− groups in each T stage (all p < 0.001). Among LN+ patients, 5-year DFS declined from 84% in T1 to 77% in T2 and 69% in T3 tumors (Fig. 1c). In LN− patients, DFS rates were 92% for T1, 91% for T2, and 84% for T3 tumors (Fig. 1d).

Five-year OS and DFS also differed significantly between LN+ and LN− patients across all molecular subtypes (all p < 0.001). In the LN+ group, 5-year OS was 89% for luminal A, 87% for luminal B, 80% for HER2-enriched, and 76% for triple-negative tumors (Fig. 2a). These rates were consistently higher in LN− patients: 95%, 94%, 92%, and 90%, respectively (Fig. 2b). For DFS, LN+ patients had rates of 80% in both luminal A and B subtypes, and 70% in both HER2-enriched and triple-negative subtypes (Fig. 2c). In LN− patients, DFS rates were 91% (luminal A), 90% (luminal B), 86% (HER2-enriched), and 87% (triple-negative) (Fig. 2d).

**Figure 2.**
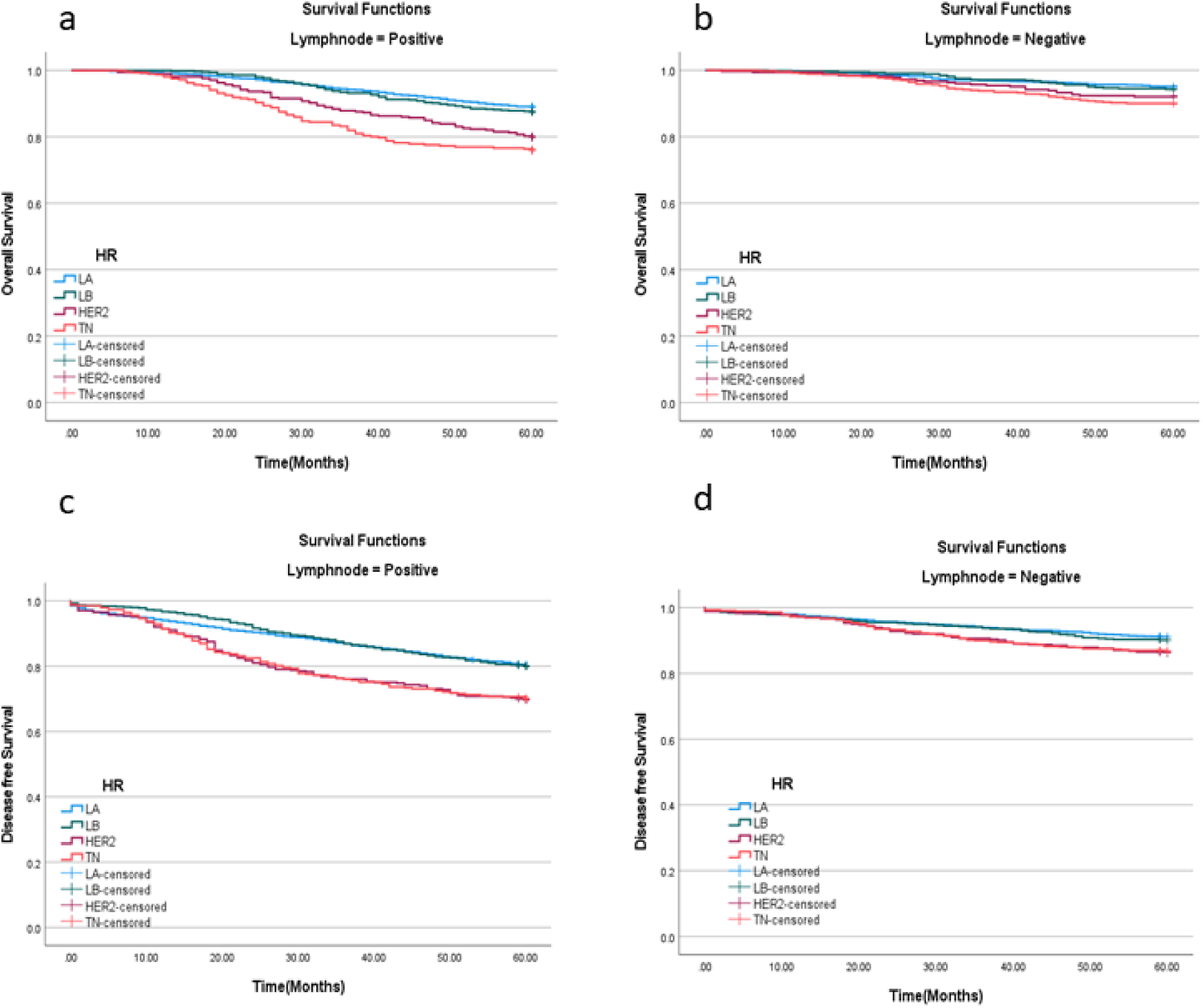
Kaplan–Meier curves for overall survival in LN+ (a) and LN− (b) patients, and disease-free survival in LN+ (c) and LN− (d) patients, stratified by molecular subtype (LA: Luminal A; LB: Luminal B; HER2: HER2-enriched

Among patients with local recurrence, 5-year OS was numerically higher in the LN− group (88% vs. 83% in LN+), but the difference was not statistically significant (p = 0.224). In contrast, among those with distant metastasis, 5-year OS was significantly lower in LN+ patients (58% vs. 62%; p = 0.013).

### Survival Analyses in Subgroups by Molecular Subtype

Table 4 presents 5-year OS and DFS according to tumor size, lymph node status, and molecular subtype.

**Table 4.** Five-year overall survival (OS) and disease-free survival (DFS) across simultaneous categories of tumor size, lymph node status, and molecular subtype.

|  |  | Molecular subtypes |  |  |  |  |
| --- | --- | --- | --- | --- | --- | --- |
|  |  | Luminal A | Luminal B | Her2-enriched | Triple negative | P Value |
|  |  | Overall survival |  |  |  |  |
| Tumor size | Lymph node status |  |  |  |  |  |
| T1 (≤2 cm) | Positive | 93% | 90% | 79% | 86% | 0.001 |
|  | Negative | 97% | 94% | 94% | 90% | 0.20 |
| T2 (2-5 cm) | Positive | 89% | 88% | 83% | 76% | < 0.001 |
|  | Negative | 95% | 96% | 93% | 91% | 0.009 |
| T3 (>5cm) | Positive | 79% | 74% | 78% | 70% | 0.563 |
|  | Negative | 92% | 95% | 92% | 74% | 0.022 |
|  |  | Diseae free survival |  |  |  |  |
| Tumor size | Lymph node status |  |  |  |  |  |
| T1 (≤2 cm) | Positive | 88% | 85% | 71% | 76% | < 0.001 |
|  | Negative | 94% | 91% | 90% | 86% | 0.13 |
| T2 (2-5 cm) | Positive | 79% | 79% | 73% | 71% | 0.008 |
|  | Negative | 91% | 93% | 85% | 88% | 0.19 |
| T3 (>5cm) | Positive | 70% | 64% | 63% | 59% | 0.706 |
|  | Negative | 92% | 95% | 80% | 73% | 0.020 |

In T1 tumors (≤2 cm), node-negative patients demonstrated excellent outcomes (OS: 90–97%; DFS: 86–94%) with no significant differences across molecular subtypes. In node-positive T1 patients, both OS and DFS varied significantly by subtype (p = 0.001 and p < 0.001, respectively), with the poorest outcomes observed in HER2-enriched tumors (OS: 79%; DFS: 71%).

For T2 tumors (2–5 cm), OS differed significantly by molecular subtype in both LN+ (p < 0.001) and LN− patients (p = 0.009). While OS remained above 90% across all subtypes in node-negative cases, it declined to 76% in node-positive triple-negative tumors. DFS differed significantly only in node-positive patients (p = 0.008), ranging from 71% to 79%.

In T3 tumors (>5 cm), node-positive patients had generally poor outcomes (OS: 70–79%; DFS: 59–70%) with no significant differences across subtypes. Among node-negative T3 patients, however, survival varied significantly by molecular subtype: 5-year OS ranged from 74% (triple-negative) to 95% (luminal B) (p = 0.022), and DFS ranged from 73% to 95% (p = 0.020).

### Univariate and Multivariate Survival Analyses of Tumor and Treatment Related Factors

Variables included in the univariable Cox regression model are listed in Table 5. Breast laterality, multifocality, in situ component, and surgical margin status were not significantly associated with OS and were thus excluded from further analysis. Variables with significant associations in univariable analysis including tumor size, type of surgery, lymph node status, molecular subtype, tumor grade, neoadjuvant chemotherapy, intraoperative radiotherapy (IORT), adjuvant chemotherapy, radiotherapy, and hormone therapy, were entered into the multivariable Cox model. After adjustment, tumor grade, neoadjuvant chemotherapy, and IORT lost their significance and were excluded from the final model, presented in Table 6.

**Table 5.** Univariable Cox Regression Analysis for Overall Survival (OS)

| Characteristics |  | Univariable HR (95% CI) | P Value |
| --- | --- | --- | --- |
| Breast laterality | Right | Reference | -- |
|  | Left | 1.064(0.931 to 1.216) | 0.362 |
| Type of surgery | BCS | Reference | -- |
|  | Mastectomy | 2.177(1.882 to 2.518) | < 0.001 |
| Tumor size | T1(≤2 cm) | Reference | -- |
|  | T2(2-5 cm) | 1.303 (1.076 to 1.579) | 0.007 |
|  | T3(>5cm) | 2.710 (2.120 to 3.464) | < 0.001 |
| Tumor grade | I | Reference | -- |
|  | II | 1.274 (1.016 to 1.597) | 0.036 |
|  | III | 1.950 (1.526 to 2.491) | < 0.001 |
| Lymphovascular invasion (LVI) | No | Reference | -- |
|  | Yes | 1.500 (1.247 to 1.804) | < 0.001 |
| Lymph node status | Negative | Reference | -- |
|  | Positive | 2.427 (2.025 to 2.910) | < 0.001 |
| Molecular subtypes | Luminal A | Reference | -- |
|  | Luminal B | 1.135 (0.913 to 1.411) | 0.255 |
|  | Her2-enriched | 1.837 (1.468 to 2.297) | < 0.001 |
|  | Triple negative | 2.283 (1.858 to 2.806) | < 0.001 |
| Multifocality | No | Reference | -- |
|  | Yes | 0.739 (0.531 to 1.028) | 0.072 |
| In situ component | No | Reference | -- |
|  | Yes | 0.881 (0.771 to 1.008) | 0.065 |
| Tumor necrosis | No | Reference | -- |
|  | Yes | 1.104 (1.052 to 1.263) | 0.034 |
| Surgical margin status | Free | Reference | -- |
|  | Close | 0.852 (0.120 to 6.059) | 0.873 |
|  | Positive | 0.924 (0.599 to 1.425) | 0.720 |
| Neoadjuvant chemotherapy | No | Reference | -- |
|  | Yes | 1.232 (1.032 to 1.471) | 0.021 |
| Adjuvant chemotherapy | No | Reference | -- |
|  | Yes | 0.343 (0.299 to 0.394) | < 0.001 |
| Intraoperative Radiotherapy (IORT) | No | Reference | -- |
|  | Yes | 0.283 (0.141 to 0.569) | < 0.001 |
| Radiotherapy | No | Reference | -- |
|  | Yes | 0.455 (0.396 to 0.523) | < 0.001 |
| Hormone therapy | No | Reference | -- |
|  | Yes | 0.361 (0.316 to 0.413) | < 0.001 |

**Table 6.** Multivariable Cox Regression Analysis for Overall Survival (OS)

| Characteristics |  | Multivariable HR (95% CI) | P Value |
| --- | --- | --- | --- |
| Type of surgery | BCS | Reference | -- |
|  | Mastectomy | 1.323 (1.057 to 1.657) | < 0.001 |
| Tumor size | T1(≤2 cm) | Reference | -- |
|  | T2(2-5 cm) | 1.304 (1.040 to 1.635) | 0.021 |
|  | T3(>5cm) | 2.021 (1.503 to 2.719) | < 0.001 |
| Tumor necrosis | No | Reference | -- |
|  | Yes | 1.142 (1.011 to 1.170) | 0.049 |
| Lymphovascular invasion (LVI) | No | Reference | -- |
|  | Yes | 1.323 (1.057 to 1.657) | < 0.001 |
| Lymp node status | Negative | Reference | -- |
|  | Positive | 2.351 (1.919 to 2.880) | < 0.001 |
| Molecular subtypes | Luminal A | Reference | -- |
|  | Luminal B | 1.118 (0.880 to 1.421) | 0.362 |
|  | Her2-enriched | 1.634 (1.265 to 2.109) | < 0.001 |
|  | Triple negative | 2.388 (1.897 to 3.006) | < 0.001 |
| Adjuvant chemotherapy | No | Reference | -- |
|  | Yes | 0.527 (0.253 to 0.689) | < 0.001 |
| Radiotherapy | No | Reference | -- |
|  | Yes | 0.624 (0.505 to 0.772) | < 0.001 |
| Hormone therapy | No | Reference | -- |
|  | Yes | 0.463 (0.361 to 0.593) | < 0.001 |

In the final model, the strongest negative prognostic factors were lymph node involvement (HR 2.35, 95% CI 1.92–2.88), triple-negative subtype compared to luminal A (HR 2.39, 95% CI 1.90–3.01), and larger tumor size (T3 vs. T1; HR 2.02, 95% CI 1.50–2.72), each associated with more than a twofold increase in the risk of death over the follow-up period. Conversely, adjuvant chemotherapy, radiotherapy, and hormone therapy significantly improved OS, reducing the risk of death during follow-up by 47% (HR 0.53, 95% CI 0.25–0.69), 38% (HR 0.62, 95% CI 0.51–0.77), and 54% (HR 0.46, 95% CI 0.36–0.59), respectively.

### Construction and Validation of the Nomogram

A prognostic nomogram was developed based on the final multivariable Cox model to estimate 3-and 5-year OS (Fig.3). The total score for each patient was calculated by summing the individual point values assigned to relevant prognostic factors. Higher scores indicated a poorer prognosis. Based on the nomogram, the absence of adjuvant chemotherapy, positive lymph node status, and undergoing mastectomy had the greatest negative impact on survival. In contrast, lower scores were observed among patients with luminal A tumors, tumor size less than 2 cm, and those who received chemotherapy, radiotherapy, or hormone therapy. The total score was then mapped to corresponding probabilities of 3- and 5-year overall survival using the nomogram scale.

**Figure 3.**
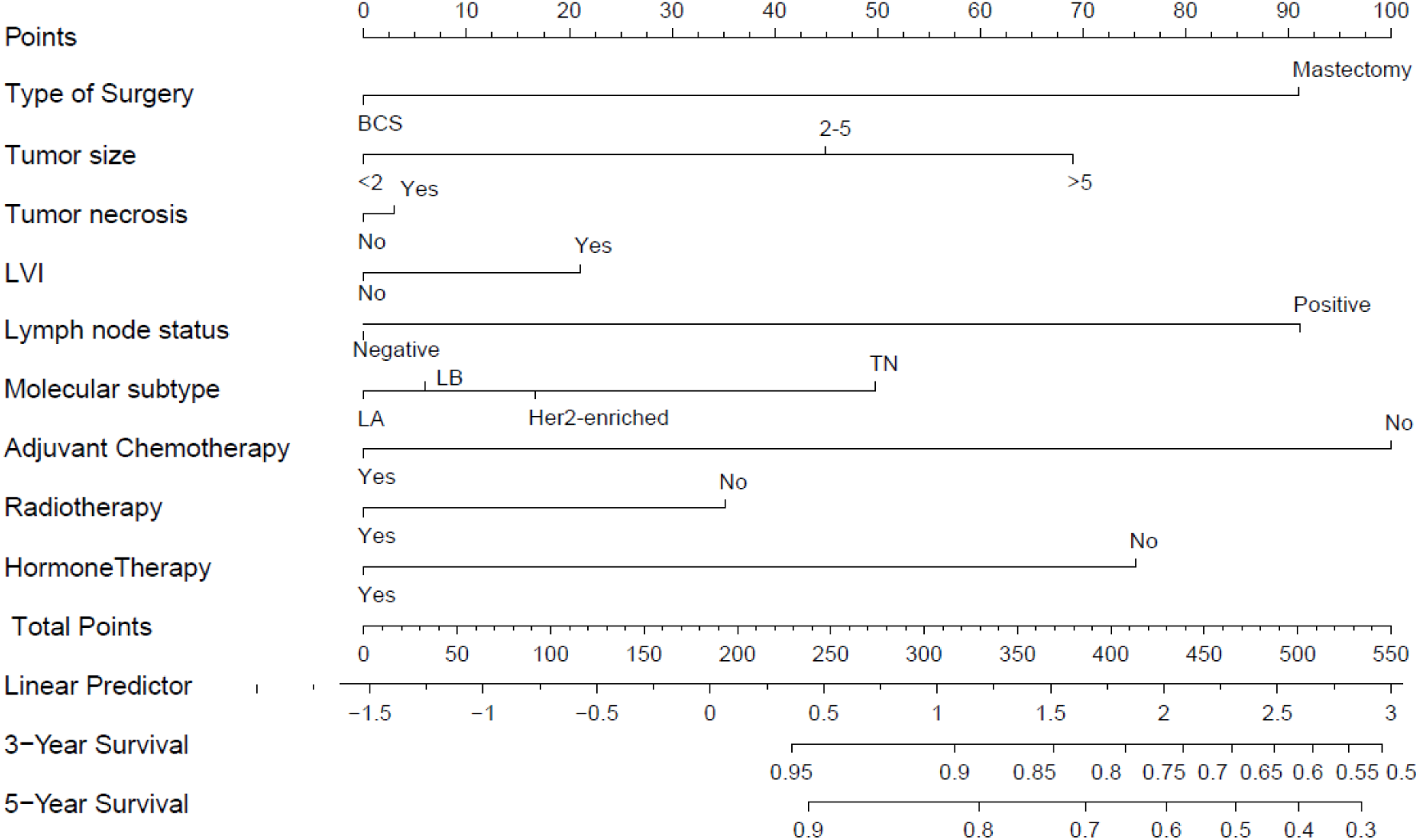
Nomogram for 3 and 5 year OS

The nomogram demonstrated good discriminative performance, with time-dependent area under the curve (AUC) values of 0.749 and 0.751 at 3- and 5-year OS, respectively (Figure 4A). Calibration plots showed close agreement between predicted and observed OS, with curves aligning closely with the 45-degree reference line (Figures 4B and 4C). The time-dependent concordance index (C-index) remained stable over the 5-year follow-up, ranging from approximately 0.70 to 0.85. These results suggest that the nomogram provides reasonably accurate survival predictions over time.

**Figure 4.**
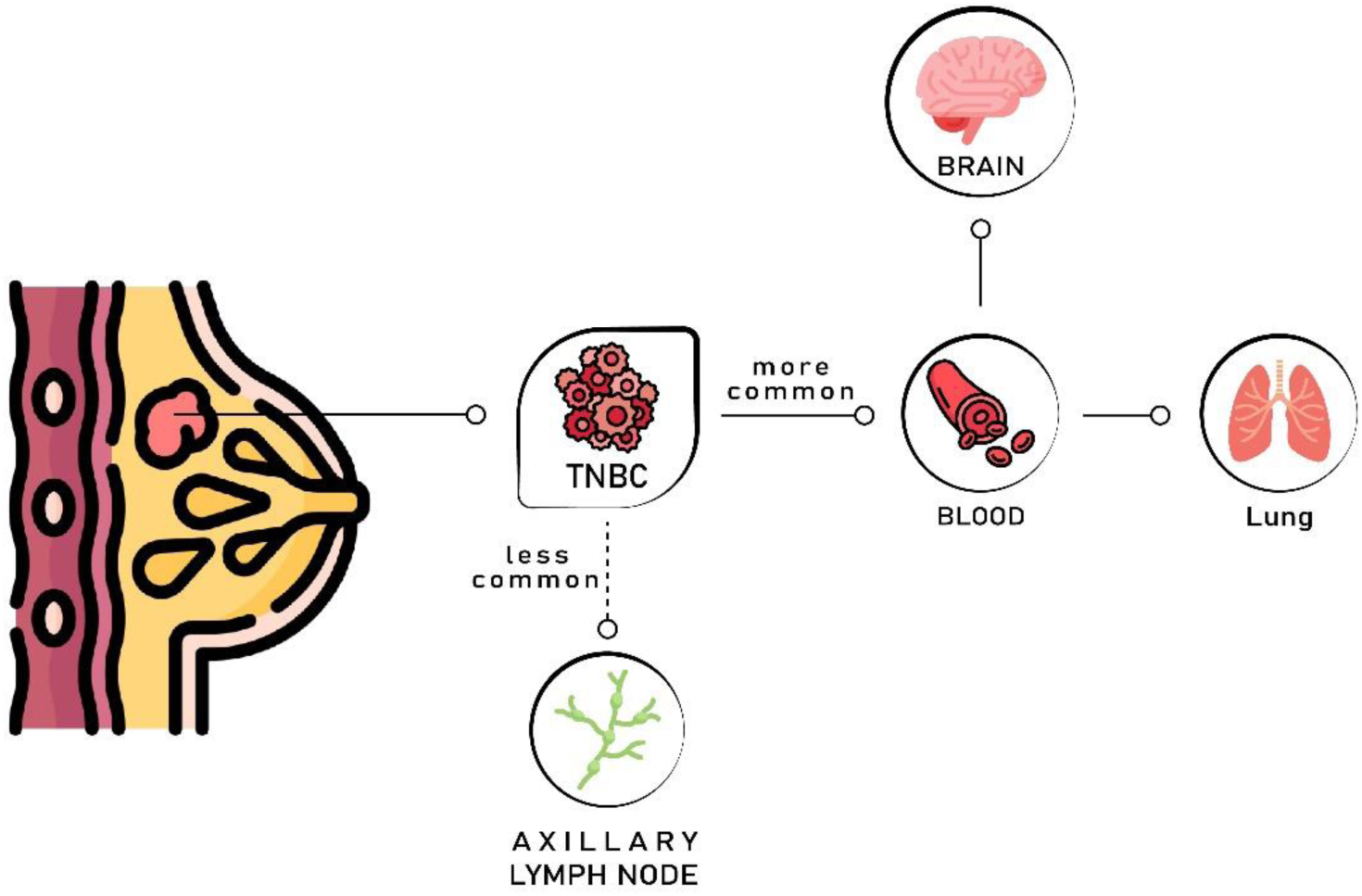
TNBC usually spreads through blood rather than lymph nodes.

## Discussion

In this study, survival analyses were conducted across subgroups defined by tumor size, lymph node involvement, and molecular subtype in the largest breast cancer cohort from southern Iran, comprising 12,225 patients. Lymph node positive tumors were significantly associated with poorer overall survival and disease-free survival across nearly all tumor size categories and molecular subtypes. The survival difference between lymph node-positive and lymph node-negative patients was substantially greater among those with larger tumors and in the HER2-positive and TNBC subtypes.

In the overall dataset, TNBC was associated with the lowest survival, followed by HER2-positive, Luminal B, and Luminal A subtypes. However, comparisons of survival across molecular subtypes did not show statistically significant differences within every tumor size and lymph node status subgroup. In those groups where significant survival differences were observed, Luminal A and Luminal B subtypes consistently showed higher survival rates than HER2-positive and triple-negative subtypes. In contrast, survival differences between Luminal A and Luminal B, and between HER2-enriched and triple-negative subtypes, were not consistent across subgroups.

The present findings are consistent with data from the Surveillance, Epidemiology, and End Results (SEER) program in the United States between 2013 and 2019. Patients with localized breast cancer demonstrated a 5-year relative survival rate exceeding 99%. Survival declined to approximately 86% in regional disease and further decreased to 31% in metastatic disease. Among molecular subtypes, Luminal HR+/Her2- and HR+/Her2+ tumors showed the most favorable outcomes. Survival was lower in HER2-enriched (HR-/Her2+) tumors, whereas triple-negative breast cancer demonstrated the poorest prognosis, with a 5-year relative survival of 77.6% (15). Teng et al. conducted survival analyses using SEER breast cancer data, including 279,078 patients diagnosed between 2004 and 2015. Based on their findings, the N0 (no nodal metastasis) and ITC (isolated tumor cells < 0.2 mm) groups showed almost identical survival curves. No statistically significant differences were observed between these groups in either overall survival (OS) or breast cancer-specific survival (BCSS). Patients with micrometastases (0.2–2.0 mm) had slightly but significantly worse survival than both the N0 and ITC groups (p < 0.001 for both OS and BCSS). In addition, patients with metastasis to the internal mammary lymph nodes had a significantly worse prognosis than all other groups, including those with axillary lymph node metastasis (16). Similarly, de Boer et al. reported that even minimal nodal involvement, including isolated tumor cells (ITCs) and micrometastases, was associated with significantly poorer survival outcomes compared with node-negative disease when adjuvant therapy was not administered. However, survival rates became more similar to those of LN-negative patients when patients received adjuvant therapy (17).

Not only is the extent of lymph node involvement an important determinant of patient survival, but the surgical management of lymph nodes also plays a critical prognostic role. The positive lymph node ratio (LNR) is defined as the proportion of positive lymph nodes among all excised lymph nodes. LNR and the negative lymph node (NLN) count are recognized as important prognostic factors in invasive BC surgery (18). A recent systematic review of fourteen retrospective cohort studies including 36,576 BC patients demonstrated that patients with at least 10 negative lymph nodes removed had significantly better outcomes than those with fewer than 10 NLNs. A higher NLN count was associated with improved 5-year overall survival (HR = 0.82, 95% CI: 0.74–0.90) as well as improved recurrence-free survival (HR = 0.76, 95% CI: 0.67–0.86) (19).

We evaluated survival outcomes of breast cancer molecular subtypes. Although molecular subtype generally influenced prognosis, survival differences between subtypes were not statistically significant in some subgroups stratified by lymph node status and tumor size. Previous studies have reported that Luminal A tumors have the most favorable prognosis, whereas triple-negative breast cancer exhibits the worst outcomes across subgroups. However, our findings showed some heterogeneity. In certain groups, HER2-positive tumors demonstrated the poorest survival rather than TNBC. In addition, the most favorable outcomes were observed not only in Luminal A but also occasionally in Luminal B. Nevertheless, our overall findings remained consistent with the literature, as in the overall dataset, TNBC was associated with the poorest survival outcomes, followed in ascending order by the HER2-positive, Luminal B, and Luminal A subtypes (15, 20, 21).

Logistic regression analysis was used to evaluate factors associated with lymph node involvement. Despite being considered the most aggressive molecular subtype of breast cancer, TNBC was associated with a significantly lower likelihood of lymph node involvement (OR = 0.54, 95% CI: 0.46–0.63). This finding is in agreement with previous studies showing that TNBC generally exhibits lower rates of nodal involvement than HER2-positive and luminal subtypes (22, 23). Si et al. analyzed 814 patients and reported results consistent with our findings. They demonstrated a positive correlation between tumor size and lymph node involvement. In addition, TNBC most frequently presents as node-negative disease (22). Similarly, Liu et al. reported that TNBC and Luminal A tumors were predominantly node negative, whereas Luminal B and HER2-positive tumors were more frequently associated with lymph node involvement. However, their analysis did not demonstrate significant survival differences between LN-positive and LN-negative patients with TNBC or Luminal A disease. In contrast, our cohort showed that lymph node status remained a significant prognostic factor across all molecular subtypes and tumor size categories. Nevertheless, consistent with our findings, although molecular subtype generally functions as an independent prognostic factor, survival differences between subtypes were not significant in certain subgroups (23).

TNBC tends to favor hematogenous dissemination rather than lymphatic spread. It metastasizes less frequently to axillary lymph nodes and bones than non-TNBC tumors do. Instead, it shows a strong preference for blood-borne routes. This pattern commonly leads to metastases in the lungs and brain (fig 4) (24, 25). As indicated, although TNBC shows less frequent lymphatic spread, the survival difference between LN+ and LN-patients is much larger in TNBC compared with other molecular subtypes and even other clinicopathological variables in this study.

The prognostic significance of multiple tumor-related and treatment-related variables was evaluated in the dataset. In both univariate and multivariate analyses, tumor size, tumor necrosis, lymphovascular invasion, lymph node status, molecular subtype, type of surgery, radiotherapy, hormone therapy, and adjuvant chemotherapy remained independent prognostic factors for breast cancer prognosis. We developed and validated a nomogram based on important clinicopathological and therapeutic factors. The model showed acceptable discriminatory power, achieving a time-dependent AUC of about 0.75. It also had good calibration when predicting 3- and 5-year overall survival. Overall, it serves as a practical tool that supports individualized risk assessment.

## Limitations

Although the Shiraz Breast Cancer Registry is one of the largest single-center breast cancer databases in southern Iran and includes patients from diverse populations, countries, and ethnic backgrounds, the single-center design limits full generalizability of the findings to the broader Iranian population or to other healthcare settings. In addition, the 24-year study period from 2000 to 2024 encompassed important advances in breast cancer management that have contributed to improved patient survival; therefore, the survival estimates reported here may not fully reflect outcomes currently achievable with contemporary treatment approaches. Furthermore, only binary lymph node status was analyzed, while detailed information on the total number of lymph nodes examined and the lymph node ratio was unavailable, limiting a more precise evaluation of nodal disease burden.

## Conclusions and Future Directions

Key insights from this Southern Iranian cohort include the consistent adverse impact of nodal positivity even in otherwise favorable subgroups, the relatively lower nodal involvement in triple-negative tumors despite their aggressive behavior, and the sustained benefit of multimodal adjuvant therapies in a real-world Middle Eastern population. Future studies with detailed axillary staging and integration of emerging biomarkers are warranted to enhance precision in prognostication.

## Data Availability

The data supporting the findings of this study are available from the corresponding author upon reasonable request.
m

## Declarations

### Ethics approval

The proposal of this study, which was reviewed and approved by Shiraz University of Medical Sciences’ ethical committee (IR.SUMS.REC.1404.370), met the highest ethical standards. All study protocols followed the guidelines stated in the Declaration of Helsinki.

### Consent to participate

Written and informed consent was obtained from the patients for participation in this study.

### Consent to publication

Not applicable

### Author Contributions

Masoumeh Ghoddusi Johari and Majid Akrami assisted with study design and data interpretation. Zahra Keumarsi conducted the data analysis. Hooman Arianpour, Nastaran Tavakolian, and Amirhessam Moosazadeh drafted the manuscript. Majid Akrami, Masoumeh Ghoddusi Johari, Vahid Zangouri, and Abdolrasoul Talei critically reviewed the final draft. All authors approved the final manuscript.

### Funding

Not applicable

### Competing Interests

The authors declare that they have no competing financial or non-financial interests relevant to the content of this article.

## Acknowledgements

Not applicable

## Notes

### Competing Interest Statement

The authors have declared no competing interest.

